# An Evidence-based Assessment of Genes in Dilated Cardiomyopathy

**DOI:** 10.1101/2020.12.10.20247197

**Authors:** Elizabeth Jordan, Laiken Peterson, Tomohiko Ai, Babken Asatryan, Lucas Bronicki, Emily Brown, Rudy Celeghin, Matthew Edwards, Judy Fan, Jodie Ingles, Cynthia A. James, Olga Jarinova, Renee Johnson, Daniel P. Judge, Najim Lahrouchi, Ronald Lekanne Deprez, R. Thomas Lumbers, Francesco Mazzarotto, Argelia Medeiros Domingo, Rebecca Miller, Ana Morales, Brittney Murray, Stacey Peters, Kalliopi Pilichou, Alexandros Protonotarios, Christopher Semsarian, Palak Shah, Petros Syrris, Courtney Thaxton, J. Peter van Tintelen, Roddy Walsh, Jessica Wang, James Ware, Ray E. Hershberger

## Abstract

**Background:** The cardiomyopathies are classically categorized as hypertrophic (HCM), dilated (DCM), and arrhythmogenic right ventricular (ARVC), and each have a signature genetic theme. HCM and ARVC are largely understood as genetic diseases of sarcomere or desmosome proteins, respectively. In contrast, >250 genes spanning more than 10 gene ontologies have been implicated in DCM, representing a complex and diverse genetic architecture. To clarify this, a systematic curation of evidence to establish the relationship of genes with DCM was conducted.

**Methods:** An international Panel with clinical and scientific expertise in DCM genetics was assembled to evaluate evidence supporting monogenic relationships of genes with idiopathic DCM. The Panel utilized the ClinGen semi-quantitative gene-disease clinical validity classification framework.

**Results:** Fifty-one genes with human genetic evidence were curated. Twelve genes (23%) from eight gene ontologies were classified as having definitive (*BAG3, DES, FLNC, LMNA, MYH7, PLN, RBM20, SCN5A, TNNC1, TNNT2, TTN*) or strong (*DSP*) evidence. Seven genes (14%) (*ACTC1, ACTN2, JPH2, NEXN, TNNI3, TPM1, VCL*) including two additional ontologies were classified as moderate evidence; these genes are likely to emerge as strong or definitive with additional evidence. Of the 19 genes classified as definitive, strong or moderate, six were similarly classified for HCM and three for ARVC. Of the remaining 32 genes (63%), 25 (49%) had limited evidence, 4 (8%) were disputed, 2 (4%) had no disease relationship, and 1 (2%) was supported by animal model data only. Of 16 commercially available genetic testing panels evaluated, most definitive genes were included, but panels also included numerous genes with minimal human evidence.

**Conclusions:** In a systematic curation of published evidence for genes considered relevant for monogenic DCM, 12 were classified as definitive or strong and seven as moderate evidence spanning 10 gene ontologies. Notably, these 19 genes only explain a minority of DCM cases, leaving the remainder of DCM genetic architecture incompletely addressed. While clinical genetic testing panels include most high evidence genes, genes lacking robust evidence are also commonly included. Until the genetic architecture of DCM is more fully defined, care should be taken in the interpretation of variable evidence DCM genes in clinical practice.

## INTRODUCTION

The major cardiomyopathies, diseases of the myocardium, have been clinically classified as hypertrophic (HCM), dilated (DCM), and arrhythmogenic right ventricular cardiomyopathy (ARVC).^1^ Each has been defined by ventricular structure and function, and supplemented in the case of ARVC by arrhythmia data. Large families with HCM,^2^ DCM^3^, and ARVC^4^ provided the basis for discovery of the first genes harboring variants causing these phenotypes.^5-7^ Based on further extensive genetic investigations, HCM and ARVC are now well-established as predominantly diseases of genes encoding key proteins of the sarcomere^8^ or desmosome,^9^ respectively.

In contrast to the genetic themes observed in HCM and ARVC, DCM has a diverse genetic architecture spanning more than ten gene ontologies.^10^ The ultimate explanation for this diversity of genetic architecture in the development of DCM remains incompletely understood, but in large relief, DCM may be considered an end- or final phenotype^11^ that occurs when cellular pathways maintaining force of contraction or ventricular structural integrity become disrupted by pathologic variation of genes encoding key proteins.

The number of genes suggested to be relevant for DCM has grown to be very large, in part due to this diverse architecture. Accepting the thesis that DCM is an end- or final phenotype, and one resulting from a myriad of possible structural, physiologic or metabolic pathway derangements, DCM candidate genes rightfully number in the hundreds. More broadly, the effort to establish a causal relationship between sequence variants in a gene and a disease is a critical step not only for cardiovascular research, but for the translation of clinical genetics to patient and family-based care.

The National Institute of Health (NIH) Clinical Genome Resource (ClinGen)^12^ has provided a semi-quantitative method to assess the clinical validity of gene-disease relationships.^13^ A Panel, composed of cardiologists, genetic counselors, and genetics and laboratory scientists with relevant expertise, applied this method to published evidence in DCM, one implemented by other ClinGen cardiovascular domain gene curation Panels, including HCM,^14^ ARVC,^15^ thoracic aortic aneurysm,^16^ and the Long QT^17^ and Brugada syndromes.^18^ Here we report the results of the evidence-based appraisal of genes associated with DCM and the implications of these findings.

## METHODS

An international group of individuals from diverse clinical and scientific backgrounds relevant to DCM was assembled as a DCM Gene Curation Expert Panel to implement the ClinGen gene-disease clinical validity classification standards^13^ with specifications to DCM. An initial set of 267 genes was identified from a structured literature search and from gene-disease reference resources (Supplementary Material). This initial list was triaged to 56 in order to remove genes that were associated with syndromes or other cardiovascular disease(s), had no direct human relevance, or represented candidate genes. The ClinGen precuration process was performed to establish the relevance for DCM, resulting in a final set of 51 genes proposed to have a monogenic role in isolated, idiopathic DCM in humans (Table 1). Additional details regarding Panel membership, operational implementation, and development of the gene list can be found in the data supplement (Supplementary Material). The data that support the findings of this study are published on the ClinGen website (https://clinicalgenome.org/). No institutional review board approval was required for this work.

**Table 1.** Quantitative scores and final classifications of genes curated for DCM. The quantitative genetic and experimental evidence points assigned for each gene and the mode of inheritance evidence is shown. MOI=Mode of Inheritance; AD=Autosomal Dominant; AR= Autosomal Recessive; SD= Semi-Dominant. Genes are sorted alphabetically and by classification category. For scores noted with an asterisk, the final assigned classification differs from the quantitative classification due to judgement of the gene curation Panel. A link to the curation summary to each corresponding final classification scoring and summary for each gene on clinicalgenome.org is provided in the classification column.

| Gene | Protein | MOI | Genetic Evidence | Experimental Evidence | Total Score | Classification |
| --- | --- | --- | --- | --- | --- | --- |
| *BAG3* | BCL2-associated athanogene 3 | AD | 12 | 6 | 18 | Definitive |
| *DES* | Desmin | AD | 7 | 6 | 13 | Definitive |
| *FLNC* | Filamin C | AD | 12 | 4 | 16 | Definitive |
| *LMNA* | Lamin A/C | AD | 12 | 6 | 18 | Definitive |
| *MYH7* | Myosin heavy chain 7 | AD | 12 | 4 | 16 | Definitive |
| *PLN* | Phospholamban | AD | 10.5 | 6 | 16.5 | Definitive |
| *RBM20* | RNA-binding motif protein 20 | AD | 11.5 | 6 | 17.5 | Definitive |
| *SCN5A* | Sodium voltage-gated channel, alpha subunit 5 | AD | 11.6 | 4.5 | 16.1 | Definitive |
| *TNNC1* | Troponin C | AD | 10.3 | 5.5 | 15.8 | Definitive |
| *TNNT2* | Troponin T2 | AD | 12 | 6 | 18 | Definitive |
| *TTN* | Titin | AD | 12 | 6 | 18 | Definitive |
| *DSP* | Desmoplakin | AD | 12 | 1.5 | 13.5 | Strong |
| *ACTC1* | Alpha actin | AD | 1.9 | 6 | 7.9 | Moderate |
| *ACTN2* | Actinin alpha 2 | AD | 4.6 | 4.5 | 9.1 | Moderate |
| *JPH2* | Junctophilin 2 | SD | 5 | 5 | 10 | Moderate |
| *NEXN* | Nexilin F-actin-binding protein | AD | 3.95 | 6 | 9.95 | Moderate |
| *TNNI3* | Troponin I | AD | 6 | 4 | 10 | Moderate |
| *TPM1* | Tropomyosin 1 | AD | 5.5 | 5.5 | 11 | Moderate |
| *VCL* | Vinculin | AD | 7.45 | 2 | 9.45 | Moderate |
| *ABCC9* | ATP-binding cassette, subfamily C, member 9 | AD | 1.5 | 2.5 | 4 | Limited |
| *ANKRD1* | Ankryn repeat domain-containing protein 1 | AD | 1.7 | 1.5 | 3.2 | Limited |
| *CSRP3* | Cysteine and glycine rich protein 3 | AD | 0.2 | 4 | 4.1 | Limited |
| *CTF1* | Cardiotrophin 1 | AD | 0.3 | 2.5 | 2.8 | Limited |
| *DSG2* | Desmoglein 2 | AD | 1.5 | 1 | 2.5 | Limited |
| *DTNA* | Dystrobrevin, Alpha | AD | 0.3 | 2 | 2.3 | Limited |
| *EYA4* | EYA transcriptional coactivator and phosphatase 4 | AD | 2.5 | 2.5 | 5 | Limited |
| *GATAD1* | Gata zinc finger domain-containing protein 1 | AR | 3 | 2.5 | 5.5 | Limited |
| *ILK* | Integrin-linked kinase | AD | 1 | 4 | 5 | Limited |
| *LAMA4* | Laminin, Alpha 4 | AD | 1.5 | 2 | 3.5 | Limited |
| *LDB3* | LIM domain-binding 3 | AD | 2.65 | 6 | 8.65* | Limited |
| *MYBPC3* | Myosin-binding protein C | AD | 6.05 | 2 | 8.05* | Limited |
| *MYH6* | Myosin heavy chain 6 | AD | 2.5 | 2 | 4.5 | Limited |
| *MYL2* | Myosin light chain 2 | AD | 0.5 | 3.5 | 4 | Limited |
| *MYPN* | Myopalladin | AD | 2.15 | 2.5 | 4.65 | Limited |
| *NEBL* | Nebulette | AD | 0.25 | 2 | 2.25 | Limited |
| *NKX2-5* | Nexilin F-actin-binding protein | AD | 1.2 | 3 | 4.2 | Limited |
| *OBSCN* | Obscurin | AD | 2 | 1.5 | 3.5 | Limited |
| *PLEKHM2* | Pleckstrin homology domain-containing protein, family M, member 2 | AR | 1.5 | 0.5 | 2 | Limited |
| *PRDM16* | PR domain-containing protein 16 | AD | 3.5 | 2.5 | 6 | Limited |
| *PSEN2* | Presenilin 2 | AD | 1.2 | 1 | 2.2 | Limited |
| *SGCD* | Sarcoglycan, delta | AD | 0.1 | 4.5 | 4.6 | Limited |
| *TBX20* | T-box transcription factor 20 | AD | 1.2 | 3.5 | 4.7 | Limited |
| *TCAP* | Titin-cap | AD | 2.4 | 1.5 | 3.9 | Limited |
| *TNNI3K* | TNNI3-interacting kinase | AD | 0.1 | 3.5 | 3.6 | Limited |
| *LRRC10* | Leucine-rich repeat-containing protein 10 | AR | 0 | 6 | 6 | No Known Disease Relationship-Animal Model Only |
| *NPPA* | Natriuretic peptide precursor A | AR | 0 | 0.5 | 0.5 | No Known Disease Relationship |
| *MIB1* | Mindbomb E3 ubiquitin protein ligase 1 | AD | 0 | 2.5 | 2.5 | No Known Disease Relationship |
| *MYL3* | Myosin light chain 3 | AD | 0.1 | 0 | 0.1* | Disputed |
| *PDLIM3* | PDZ and LIM domain protein 3 | AD | 0 | 0 | 0* | Disputed |
| *PKP2* | Plakophilin 2 | AD | 0.3 | 0.5 | 0.8* | Disputed |
| *PSEN1* | Presenilin 1 | AD | 0 | 0.5 | 0.5 | Disputed |

### Phenotype definition

The DCM phenotype was defined by systolic dysfunction, conventionally noted as a left ventricular (LV) ejection fraction (LVEF) of <50% accompanied by LV enlargement (LVE), after other usual clinically detectable causes of cardiomyopathy were excluded. Dilated cardiomyopathy presenting during pregnancy (peripartum or pregnancy-associated DCM (PPCM; PACM) was included as published evidence has demonstrated a genetic background in PPCM/PACM that is similar to idiopathic DCM.^19, 20^ In addition, DCM observed in conjunction with a LV non-compaction phenotype was also evaluated and contributed to evidence scores. Publications used for gene scoring were required to specify how the DCM phenotype was defined, and that other usual causes (except genetic) were excluded. In the absence of such specifications, the data were either not scored, or the score was reduced from the default points recommended by the ClinGen standard operating procedure (version 7) for the type of variant observed (https://clinicalgenome.org/curation-activities/gene-disease-validity/training-materials/).

### Gene curation and evidence scoring process

The ClinGen gene curation scoring framework^13^ sums scores for published clinical genetic and experimental laboratory evidence. Members of the Panel, trained to curate following the ClinGen protocol, scored published evidence according to the gene-disease clinical validity standard operating procedure version 7. This was presented to the full Panel on conference calls to establish an approved clinical validity classification. Genetic evidence was comprised of case-level data, including variant evidence and segregation, in addition to case-control data. Variants shown to be absent or have a minor allele frequency (MAF) of <0.0001 in gnomAD^21^ (https://gnomad.broadinstitute.org/) were evaluated and scored. Individuals or pedigrees with more than one possibly relevant variant in any putative DCM gene were not scored. Experimental evidence was assessed by category (expression data, functional alterations, model systems, and rescue). Additional details are provided in the Supplementary Material and as previously published.^13^

The ClinGen clinical validity classifications include “strong” (12-18 points), “moderate” (7-11 points), “limited” (1-6 points), and “no known disease relationship” (0 points of scorable genetic evidence). The maximum number genetic evidence points that could be given was 12, and maximum experimental 6, for a highest total possible score not exceeding 18 points. “Definitive” was defined as a gene with a strong evidence score with multiple publications over at least 3 years and no contradictory evidence. If the numeric, point-based classification was not considered to reflect the collective assessment of the Panel’s clinical and scientific experience, the classification was further discussed and, when applicable, the final classification was modified to a classification reflective of consensus of the Panel. Additional details regarding the clinical validity classification definitions and scoring system are available in the Supplementary Material and the SOP (https://clinicalgenome.org/docs/summary-of-updates-to-the-clingen-gene-clinical-validity-curation-sop-version-7/).

### Composition of clinical genetic testing panels

Sixteen commercially available clinical genetic testing panels curated for DCM were assessed for the presence or absence of the 51 genes curated herein. Panels were identified through a query of the NCBI Genetic Testing Registry^22^ by searching the term “dilated cardiomyopathy.” The final panel evaluation included targeted DCM multi-gene panels.

## RESULTS

### Summary of DCM Gene Classifications

Fifty-one genes were identified as having a role in isolated, idiopathic DCM (Table 1) as described above. The appraisal of genetic and experimental evidence resulted in 19 genes with substantial evidence supporting a role in monogenic DCM, including 11 definitive (21%), 1 strong (2%), and 7 (14%) moderate evidence classifications (Table 1; Figure 1) from ten gene ontologies (Figure 2). Notably, more than half of genes curated (63%) were determined to be of limited evidence (n=25, 49%), disputed (n=4, 8%), to have no known disease relationship (n=2, 4%), or supported by animal model data only (n=1, 2%).

**Figure 1:**
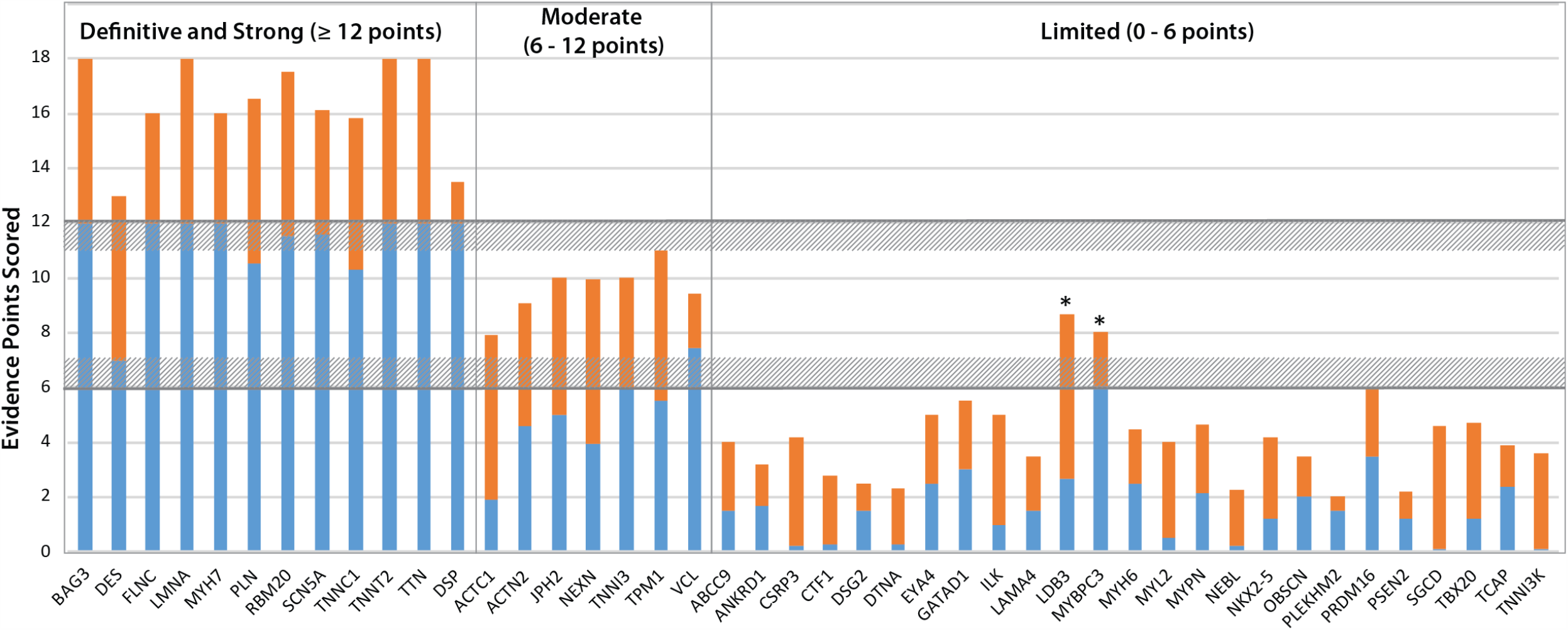
Quantitative contributions of genetic and experimental evidence to the clinical validity classifications of genes curated for DCM. The sum of genetic (blue) and experimental (orange) evidence scores are shown for genes classified as having definitive, strong, moderate or limited evidence in a monogenic relationship with DCM. The two genes noted with an asterisk had quantitative scores within the quantitative range for a moderate classification, but a limited classification was assigned at Panel review (see text).

**Figure 2.**
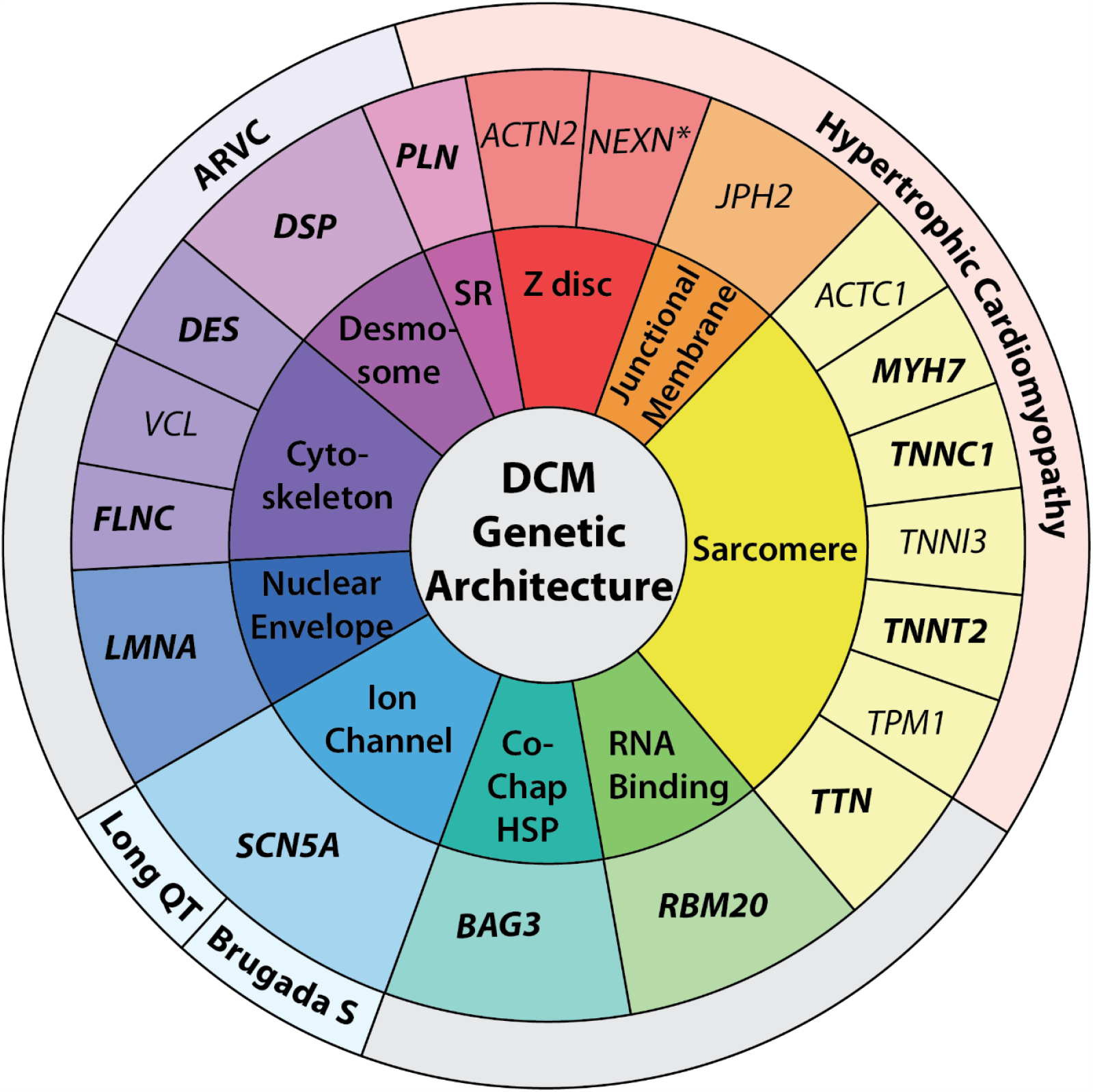
The genetic architecture of DCM. The genetic architecture of DCM spans ten gene ontologies, as shown in the innermost colored text circle. Ontology abbreviations include: SR, sarcoplasmic reticulum; Co-Chap HSP, co-chaperone, heat shock protein. The middle text circle specifies genes classified as strong or definitive (bold text) or moderate (regular text) for DCM, organized by gene ontology. Of the 19 DCM genes shown, 14 have been evaluated by other ClinGen gene curations for Hypertrophic Cardiomyopathy or Arrhythmogenic Right Ventricular Cardiomyopathy, and the channelopathies including the Long QT and Brugada Syndromes (Brugada S). Each of these genes have also been classified as moderate, strong, or definitive for these other phenotypes, except for *NEXN*, noted with an asterisk, which has been classified as having limited evidence in HCM.

### Definitive/Strong Classifications

A total of 12 genes were classified as a definitive or strong relationship (*BAG3, DES, DSP, FLNC, LMNA, MYH7, PLN, RBM20, SCN5A, TNNC1, TNNT2*, and *TTN*). By definition, strong or definitive classifications represent genes that have a role in DCM that has been clearly demonstrated in the literature over time. While the ClinGen framework requires a minimum of two independent publications to reach a strong or definitive classification, each definitive/strong gene-disease relationship had an abundance of genetic and experimental evidence, ranging from 7-12 points and 4.5-6 points, respectively, for those classified as definitive. In addition, genes that demonstrated significant enrichment for rare variants in a recently published DCM case-control analysis^23^ also emerged as strong or definitive when performing this evidence assessment.

*DSP* was the only gene with a score ≥12 points that remained classified as strong rather than definitive evidence. While the criteria for replication over time was met, and substantial genetic evidence has been published from rigorously phenotyped cohorts meeting DCM criteria and without clinical evidence of ARVC, curation of experimental evidence presented challenges in scoring due to arrhythmic phenotypes complicating the interpretation of experimental data, resulting in a low experimental evidence score of 1.5 points. The Panel ultimately elected to assign a strong classification to *DSP* with opportunities for future curation to reappraise this gene with improved clarification of the interrelationships of *DSP*-related phenotypes,, as well as other genes, where the relationship of DCM to arrhythmia adds complexity that exceeds a disciplined approach using current ClinGen curation guidance.

In the curation of *RBM20*, several variants contributing to the genetic evidence score included those within the described hotspot region in exon 9 (amino acids 634, 636, 637, and 638).^24^ However, additional missense variants were identified and scored outside of the hotspot region, providing support that variation in addition to that of the exon 9 region may also contribute to the DCM phenotype. Scored variation in *TTN* was restricted to premature termination codons, with most located in exons constitutively expressed in the adult heart and in the A-band. This was an anticipated finding that provided robust support of pathogenicity, as it has been well established that *TTN* alterations of this type are overrepresented in individuals with DCM.^25-29^ Given the complexity of *TTN* variant architecture and the broad contribution of *TTN* variation in DCM beyond the A band represented in our data curation from the published literature, future *TTN* domain- and band-specific curations integrated with expression data should be considered.

### Moderate Classifications

A total of seven genes were classified as having a moderate level of evidence, namely *ACTC1, ACTN2, JPH2, NEXN, TNNI3, TPM1*, and *VCL*. The range of genetic evidence points varied widely from 1.9-7.45, and experimental evidence varied from 2-6 points, with total summative scores ranging from 7.9-11 points. With the exceptions of 2 genes (*VCL* and *ACTC1*), the contributions of genetic and experimental data to the final classifications were generally balanced (Figure 1). In the case of *ACTC1*, a proportionally higher experimental score (6 points), with less substantial published clinical DCM data (1.9 points), contributed to the moderate classification. Conversely, *VCL* has the highest genetic evidence score in the moderate evidence genes (7.45 points), and while in vitro protein-protein assays, expression studies, and animal models have been published supporting the role of *VCL* in DCM, existing studies could only be scored for 2 points of experimental evidence. With the score of the moderate-classified genes generally totaling at the upper defined range, additional published data and subsequent curations may well result in a future reclassification of these genes to strong or definitive.^30^

### Limited and No Known Disease Relationship Classifications

The majority of genes curated were deemed to have limited or no known disease relationship. The 25 genes with limited evidence (Table 1) had genetic evidence scores ranging from 0.1-6.05 points and experimental evidence scores ranging from 0.5-6 points. Two genes, *MYBPC3* and *LDB3*, had scores that numerically would have placed them into the moderate evidence category. However, after review, the Panel decided to downgrade the clinical validity classification for both genes to limited. As candidate genes for the DCM phenotype, *LDB3* and *MYBPC3* have been sequenced many times and have only accumulated modest scores, even when considering that they have been targeted in DCM genetic studies for more than 10 years. Supporting segregation and case-control data are lacking and the scored genetic evidence was interpreted as circumstantial, additively placing them into a higher category over time regardless of the absence of strongly supportive data. Therefore, the quantitative classification exceeded the Panel’s overall assessment of the clinical relevance of these genes in idiopathic DCM. Without these genes, the range of limited evidence scores are 0.1-3.5 and 0.5-4.5 for genetic and experimental evidence, respectively.

*EYA4* was curated for DCM and hearing loss as the disease entity (MONDO:0011541). While the filtering of the initial gene list excluded genes primarily related to a syndrome impacting systems beyond that of the cardiovascular system, the hearing loss phenotype observed in *EYA4* can be quite subtle and may not be clinically apparent at the time of the genetic evaluation of DCM. Therefore, the Panel curated this gene separately for its role in the DCM phenotype, with or without hearing loss, and was classified as having limited evidence.

Genes determined to have no known disease relationship, formerly referred to as “no reported evidence,” include *LRRC10, NPPA*, and *MIB1*. This classification indicates that the gene does not have human genetic evidence suggesting a causal role in monogenic DCM. While many candidate genes with currently no known relationship with human DCM were removed during the development of the initial gene list, these genes emerged due to the degree of experimental data suggesting a role in DCM development. While these genes have experimental evidence supporting a relationship with the DCM phenotype, there was an absence of human genetic evidence meeting DCM criteria defined for this curation effort (*MIB1, NPPA*). Of note, in the curation of *LRRC10*, published experimental data^31^ included non-human model organisms and accumulated an experimental evidence score much higher (6 points) than *NPPA* (0.5 points) and *MIB1* (2.5 points). In addition, while human genetic evidence for *LRRC10* had been published, the data were excluded after Panel review. Nonetheless, due to the compelling *LRRC10* animal model evidence, an added “Animal Model Only” tag to the no known disease relationship classification was added. If human data emerges for *LRRC10*, future curation may result in reclassification.

### Disputed Classifications

The disputed classification was assigned when available evidence was insufficient and question was raised regarding relevance to a monogenic causation of DCM. Four genes were disputed after curation and Panel discussion (*MYL3, PDLIM3, PKP2*, and *PSEN1*). Each of these genes had minimal genetic evidence that was able to be scored after review, mainly due to frequency of the few variants reported in the general population exceeding the defined MAF cut point of 0.0001. Further, in a recently published case-control analysis of DCM genes, *PKP2* and *PDLIM3* variants were not enriched in cases when compared to a control population.^23^ Ultimately, the Panel concluded that the current evidence was not sufficient to support a causative, monogenic relationship with the DCM phenotype.

### Composition of DCM Genes on Clinical Genetic Testing Panels

Sixteen commercially available clinical genetic testing panels were evaluated for DCM gene inclusion (Figure 3). On average, evaluated panels contained a total of 64 genes, with the total number of genes ranging from a minimum of 37 to a maximum of 123 genes. Of all panels, 50% (n=8) offered testing for ≥75% of the genes curated for DCM. Eight out of the 11 definitive genes appeared on all panels, with *TNNC1* and *TNNT2* present on 95% and *FLNC* on 75% of panels. The observation that *FLNC* is only included on 75% of clinical testing panels may be explained by its relatively recent emergence in DCM, with a first major publication in 2017.^32^ With the exception of the moderate-classified *JPH2*, which appeared in only 25% of panels, genes classified as definitive, strong, or moderate appeared on a majority of evaluated panels (75%-100%). The presence of limited genes ranged widely (13%-100%), with *ABCC9, LDB3, MYBPC3, MYH6*, and *TCAP* present on all evaluated panels and *CTF1, PLEKHM2, PSEN2, TNNI3K*, and *OBSCN* present on the fewest (12%-25%). In addition, some disputed genes were represented more commonly than those with limited evidence, for example in the cases of *PDLIM3* and *PKP2*, which were both present on 75% of evaluated panels.

**Figure 3.**
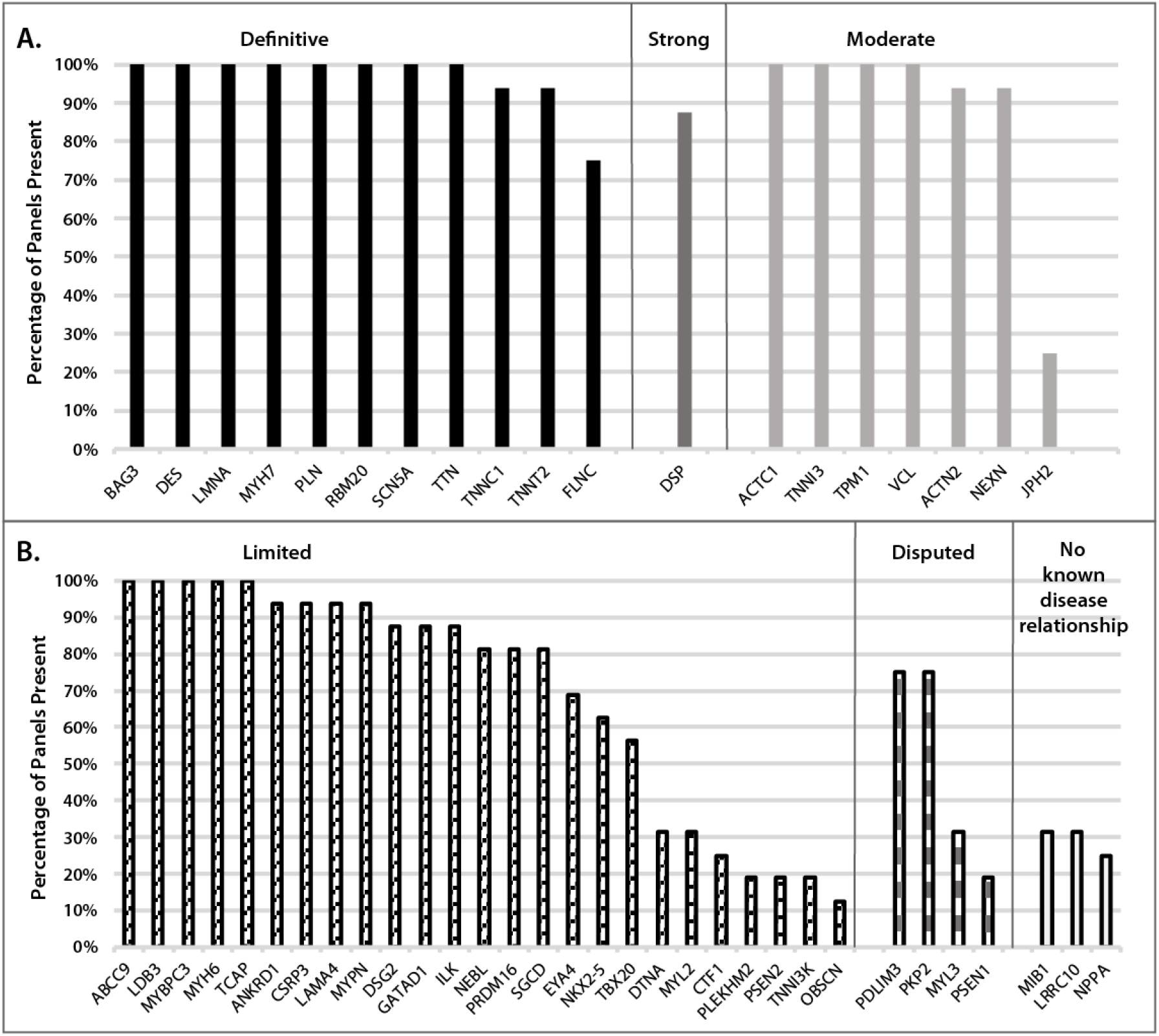
Curated genes on clinically available DCM genetic testing panels. The percentages of DCM genetic testing panels that include the genes curated for DCM herein are shown for 16 commercial laboratories identified on the NCBI genetic testing registry. Genes are grouped by clinical validity classification, ranging from definitive, strong and moderate (Panel A) to limited, disputed and no known disease relationship (Panel B).

## DISCUSSION

This study conducted a systematic review and curation of published evidence for genes considered relevant for monogenic DCM. Winnowed to 51 genes for curation from an initial list of 267 candidates, each gene was scored according to the established ClinGen framework^13^ used previously for other cardiovascular genetic conditions.^14-18^ Twelve genes were found to have definitive or strong relationships, and seven had moderate evidence for a monogenic cause of DCM. Of the remaining 32 genes, 25 were determined to be limited, and 7 were disputed or assigned as having no known disease relationship due to lack of human evidence. To our knowledge, this effort represents the first standardized curation of evidence implicated in the monogenic cause of DCM, and provides guidance to clinicians for testing strategies in the genetic evaluation of DCM.

This work underscores the diverse genetic architecture of DCM and illuminates the intersections of genes relevant for DCM with other well-established cardiovascular gene-phenotype relationships. It also illustrates the complexity of DCM genetics. Several genes curated as definitive for other cardiomyopathy or arrhythmia phenotypes were also scored as definitive for DCM (Figure 2). One of the most prominent arrhythmia genes, *SCN5A*, definitive for DCM, has also been classified as a definitive gene in Long QT Type 3^17^ and Brugada^18^ syndrome. Even though the precise molecular mechanisms that result in a DCM versus an electrophysiologic phenotype due to *SCN5A* variation remain incompletely understood, the *SCN5A* clinical and experimental evidence for DCM achieved a definitive classification. Genes encoding proteins of the desmosome have also been proposed as relevant for DCM,^33^ and *DSP, DSG2*, and *PKP2*, considered definitive for ARVC,^15^ had various degrees of evidence when curated strictly for DCM. In the case of DCM, *DSP* was scored as strong, and may likely move to a definitive classification in future re-evaluation. *DSG2* was scored as limited, and the lack of monogenic DCM evidence for *PKP2* resulted in a disputed classification. Additional human clinical genetics data from well-phenotyped cohorts will be needed to further clarify the relevance of these genes for DCM. Two sarcomere genes, *MYH7* and *TNNT2*, established as definitive for HCM, were also definitive for DCM. Three other definitive genes for HCM, *TNNI3, TPM1, ACTC1* were considered of moderate evidence for DCM, but may emerge as strong or definitive genes for DCM with additional evidence. Conversely, *TNNC1*, definitive for DCM, was scored as moderate evidence for HCM.^14^

Other genes scored as definitive with evidence principally from the DCM phenotype further highlight the diverse genetic architecture of DCM, in contrast to ARVC and HCM (Figure 2). Most notable is *TTN*, an enormous scaffolding protein of the sarcomere, which contributes the most cases of DCM.^25, 26^ For HCM and ARVC, *TTN* was classified as limited.^14^ *LMNA*, encoding a protein of the inner nuclear membrane that exhibits striking pleiotropic effects in skeletal muscle, adipose and other tissues, was considered definitive for DCM while limited for ARVC. *RBM20*, which encodes an RNA binding protein that regulates RNA splicing, was scored as definitive for DCM and has no other phenotypic representation beyond DCM. *FLNC*, an actin crosslinking protein widely expressed in cardiac and skeletal muscle, was also classified as definitive in DCM. Additional genes with moderate evidence in association with DCM also illustrate the diversity of DCM genetic architecture (Figure 2).

The work presented is an important step forward in the describing the genetic architecture of monogenic, primarily adult-onset, non-syndromic DCM. However, despite this progress, a great deal of work remains to more fully understand the genetic basis of DCM. Even after a rigorous evaluation of variants identified in DCM genes, a pathogenic or likely pathogenic classification can only be established in a minority of DCM patients, estimated at 20-35%.^34^ This modest genetic testing sensitivity is observed even in the case of multigenerational families that have multiple affected members, which on its face supports an underlying genetic cause. The reason for this low testing sensitivity even for familial DCM remains unclear. In part, this appears to be related to the already established locus heterogeneity, with a substantial number of genes already established as relevant. Notably almost all genes account for only a small percentage of cause, exceptions including *TTN* explaining up to 15-20%,^26^ *LMNA* in up to 4-6%,^35^ and *MYH7* up to about 3%^23^ of DCM cases. Whether any of the 25 genes attributed to have limited evidence will emerge as moderate, strong or definitive remains to be determined. It is also possible, if not likely, that additional genes considered novel to DCM, even from yet-to-be-included ontologies, remain to be identified.

Alternatively, genetic mechanisms exceeding those considered monogenic could well be at play. Previous evidence has suggested that some proportion of DCM may have an oligogenic or polygenic basis,^35-38^ but a DCM phenotype confounded by more than one rare variant was not accounted for under the ClinGen framework,^13^ and therefore not scored as all curations assumed a classic (monogenic) Mendelian paradigm. Based on preliminary data, this may be an important focus for ongoing effort. Other types of genetic variation that to date have received minimal investigation in DCM, including promoter variants, common variants, or structural variants exceeding in size those able to be detected by next generation sequencing, may also be relevant to define DCM genetic cause.

The findings of this curation effort are also relevant for family-based clinical genetics care of patients with DCM.^39^ Establishing genetic risk of DCM in family members as a component of a genetic evaluation presents considerable opportunity for disease mitigation and prevention. Genetic testing is a central component of a DCM genetic evaluation, and most commercially available DCM gene panels test several dozen genes, well exceeding the 19 genes curated here as definitive, strong, or moderate. In the setting of a DCM phenotype, variants reported in genes beyond these 19 can at most only be classified as variants of uncertain significance, and in the cases of disputed genes or those with no known disease relationship, identified variants are not able to reach a clinical classification. This is appropriate and nearly unavoidable, as these genes have clinical validity yet to be defined. Based on the evidence presented, moderate DCM genes may eventually gain sufficient clinical and experimental support to be assigned as strong or definitive for DCM at the time of future curation. Whether the many limited evidence genes will also accumulate sufficient clinical or experimental data over time in order to emerge as strong or definitive is remains to be determined.

As noted, one of the most significant issues for the majority of families is that with current genetic testing the genetic cause of DCM often remains elusive.^34^ In addition to marked locus heterogeneity, the modest testing sensitivity may also be explained in part by the marked allelic heterogeneity that also confounds variant interpretation, as many DCM variants are private, or even if previously observed, lack sufficient data to support pathogenicity using the current stringent standards for variant adjudication.^38, 40, 41^ Moreover, as previously mentioned, non-Mendelian mechanisms, even if clearly defined, may not be easily integrated into conventional approaches to variant interpretation.^40^

As acknowledged by other ClinGen cardiovascular domain gene curation Panels,^14-18^ clinical genetic testing panels feature many genes classified as limited, disputed, or have no known disease relationship with the phenotype of interest. Current variant adjudication guidance^38, 40, 41^ is intended for the clinical interpretation for the monogenic cause of disease in order to translate genetic test results into information to be applied to clinical, family-based care.^39^ While many genes have been suggested to have a relationship with DCM as shown in the initial expansive list, only a minority were identified to have a possible monogenic role in idiopathic DCM. The rationale for commercial sequencing panels to include genes for DCM even beyond the 44 limited, moderate, strong, and definitive evidence genes curated here is unclear, although some represent syndromic conditions where the DCM phenotype may be observed as a feature of the syndrome. It is possible that knowledge gained regarding candidate genes might benefit the research community for the purpose of discovery, or a more expansive gene list may represent other interests of commercial laboratories. Nevertheless, the inclusion of genes lacking even moderate evidence of a gene-disease relationship contributes to uncertainty in clinical care for patients and providers. This also creates the potential for misapplication of genetic information in the care for DCM patients and their at-risk family members.^39^

### Implications for Clinical Care

The results of this analysis indicate that pathogenic and likely pathogenic variants in the 19 higher evidence genes (definitive, strong, moderate) can reasonably be used for diagnostic and predictive purposes in the management of patients and families with monogenic DCM. However, it is unclear if genes assigned to limited, disputed, or no known disease relationship classifications will emerge as Mendelian causes of DCM as more evidence is published. Analysis of the currently available evidence suggests that variation in these genes is largely uninformative in isolation. With the possible exception of a large family with ample opportunity for segregation analysis, variants in genes not classified as moderate, strong, or definitive evidence will seldom be interpretable for DCM. Inclusion of such genes of uncertain significance on clinical testing panels should be applied with caution, and ideally in the context of an expert, multidisciplinary team to avoid misapplication.

## CONCLUSION

In this study, an evidence-based curation of published literature evaluating the clinical validity of the monogenic relationship with DCM was performed. Of 51 genes, 12 were classified as definitive or strong evidence and 7 moderate evidence. These 19 genes provide a solid foundation for clinical care. The remaining genes, classified as limited evidence or no known disease relationship, have limited clinical utility but may provide valuable information for investigators as additional evidence in support of genetic cause of DCM is sought. Several of the genes classified as definitive for DCM have also been classified as definitive for other distinct cardiomyopathy or arrhythmia phenotypes, underscoring the unique and diverse genetic architecture of DCM. Despite this, the current sensitivity of genetic testing in DCM of only 20-35% emphasizes the need for continued efforts to more fully understand the genetic basis of DCM, whether from known candidate genes or those not yet understood to be relevant, or from genetic mechanisms yet to be more fully described.

## Supporting information

Jordan et al Supplemental Data

## Data Availability

All data are provided in the manuscript, or the supplementary data, or the data that support the findings of this study are published on the ClinGen website (https://clinicalgenome.org/).

## ACKNOWLEDGEMENTS

We extend thanks to Stephanie Schulte, MLIS, Associate Professor and Head of Research and Education Services at the Health Sciences Library at The Ohio State University for her assistance in developing a systematic and comprehensive approach in developing the initial gene list from the OMIM, Gene, and GenBank databases.

