## Supplementary material for "An Evidence-based Assessment of Genes in Dilated Cardiomyopathy": Jordan et al Supplemental Data

### SUPPLEMENTAL MATERIAL

#### Supplementary Methods

*Panel membership and workflow*

DCM gene curation activities were carried out by an international, multi-site Panel of individuals with expertise in laboratory science, molecular diagnostics, research, and/or clinical care in idiopathic DCM. The Panel, termed "The DCM Gene Curation Expert Panel," a sub-working group of the ClinGen Cardiovascular Clinical Domain Working Group, was composed of 34 individuals from over 20 institutions representing 7 countries (United States, Canada, Netherlands, Australia, United Kingdom, Switzerland, and Italy). Using the ClinGen standard operating procedure (SOP) version 7 (https://clinicalgenome.org/docs/summary-of-updates-to-the-clingen-gene-clinical-validity-curation-sop-version-7/), curators scored available evidence and prepared a presentation for full Panel review, discussion, and approval during a biweekly conference call between September 2019 and October 2020 to establish a final classification. Curated data was entered into a web interface, the Gene Curation Interface (GCI), to organize and record evaluated evidence.

*Development of the DCM gene list*

To develop a comprehensive list of genes thought to have a role in the development of DCM in humans, a comprehensive search was conducted in three National Center for Biotechnology Information database resources in October 2017: Gene (https://www.ncbi.nlm.nih.gov/gene), Online Mendelian Inheritance in Man (OMIM)[1] (https://www.ncbi.nlm.nih.gov/omim), and GenBank[2] (https://www.ncbi.nlm.nih.gov/genbank/). Starting with Gene, the search terms "dilated cardiomyopathy" OR "cardiomyopathy, dilated" were applied and limited to Homo Sapiens to identify 199 genes. A subsequent search in OMIM and GenBank using the same search terms, limiting to Animals and Genomic DNA/RNA as the molecule type, identified a list of 204 genes. This initial list was further expanded when adding additional genes not identified in the database query from an expansive DCM gene list that had been previously published[3], which added 63 genes. The full initial list included 267 genes.

Each gene was then manually evaluated by performing a PubMed search of the "[GENE]" AND "dilated cardiomyopathy" in addition to "[GENE]" AND "cardiomyopathy, dilated" to identify relevant publications proposing a role in the development of idiopathic DCM in humans (May 2018). Genes were removed for the following reasons: those specified as possible candidate genes without supporting animal or human genetic evidence; those that did not have published evidence in human idiopathic DCM; those described as primarily part of a syndrome; and those primarily observed in other non-DCM cardiovascular diseases. Following the application of these criteria, 56 genes remained.

Genes implicated in more than one disease underwent precuration to confirm the DCM disease entity and mode of inheritance for each gene curated (https://clinicalgenome.org/working-groups/lumping-and-splitting/). Precurations were performed to identify the disease entity and mode of inheritance. Precurations were shared with the Panel for review prior to formal preparation and scoring of evidence to assign a gene-disease validity classification. During this process, five genes were removed from the curation schedule due to syndromic disease entity (*SDHA*), lack of sufficient evidence specific to the DCM phenotype as defined for this curation effort (*DSC2*, *GATA4*, *GATA6*), or a different cardiomyopathy sub-type as the primary disease entity without a clear assertion for an isolated DCM phenotype in a single gene-disease relationship (*ALPK3*). The remaining 51 genes represent the final gene list (June 2019) (Figure S1).

Five of the genes on the final list were identified as predicted to be classified as definitive (*BAG3*, *LMNA*, *MYH7*, *TNNT2*, and *TTN*) and therefore underwent an expedited curation, requiring the review and approval of two Panel members of the evidence review in an offline mechanism, followed by review and approval of the evidence summary by the full Panel prior to publishing. In addition, two genes had been recently curated by the Hypertrophic Cardiomyopathy Gene Curation Expert Panel (HCM GCEP) for an applicable disease entity, intrinsic cardiomyopathy (MONDO:0000591), and mode of inheritance prior to the DCM gene curation project (*PLN*, https://search.clinicalgenome.org/kb/gene-validity/8772; *ACTN2* https://search.clinicalgenome.org/kb/gene-validity/8772)[4]. The other 44 genes underwent the standard curation process.

*Curation of Published Evidence*

Evaluation of genetic evidence included publications presenting clinical data for patients, families, and large cohorts that could be consider for scoring of individual variants, segregation in pedigrees, and case-control analyses. A minor allele frequency (MAF) cut-off for the maximum credible frequency to have a pathogenic effect was calculated using an allele frequency calculator which integrates disease prevalence, mode of inheritance, genetic and allelic heterogeneity, and penetrance (https://www.cardiodb.org/allelefrequencyapp/).[5] We sought to produce a conservative estimate consistent with Mendelian, autosomal dominant disease, occurring at a prevalence 1/250 individuals in the population[6]. While reduced penetrance is commonly observed in DCM, penetrance estimates are not widely available. Further, the penetrance of the genes included in the curation list are anticipated to vary widely, therefore, estimates of 20-40% penetrance were considered. DCM is also highly heterogeneous, with no single variant causing more than 1-2% of DCM cases. Taking these values into consideration, a maximum credible population allele frequency of 0.0001 was generated, consistent with the clinical standard[7] for monogenic DCM. Variants with a MAF <0.0001 were scored in the curation of genetic evidence.

Experimental evidence was assessed by category (expression data, functional alterations, model systems, and rescue). Expression evidence prioritized studies of human cardiomyocytes or animal models as an alternative when human models were not available. Model systems assessing the disruption to the gene in consideration demonstrated a phenotype suggestive of DCM, and rescue models using cell-culture or non-human animal models with DCM could be rescued by restoring wild-type gene product. Experimental data only evaluating rare variants with some evidence of impact were scored.

Genes with a score reaching a strong classification that demonstrated replication over time in the literature, as defined by at least three years since original publication and at least two independent supporting publications, were classified as definitive. Limited evidence genes lack substantial evidence supporting the gene-disease relationship, however, data challenging the relationship is also not present, and therefore while the role in monogenic disease is not supported based on currently available evidence, the clinical relevance of these genes is not ruled out. Genes were classified as disputed when evidence was deemed insufficient and panel opinion and/or literature questioned the biological relevance of the gene in the disease. Genes classified as no known disease relationship and animal model only represent candidate genes that do not yet have sufficient human data to evaluate for a clinical classification.

**Supplemental Figure**

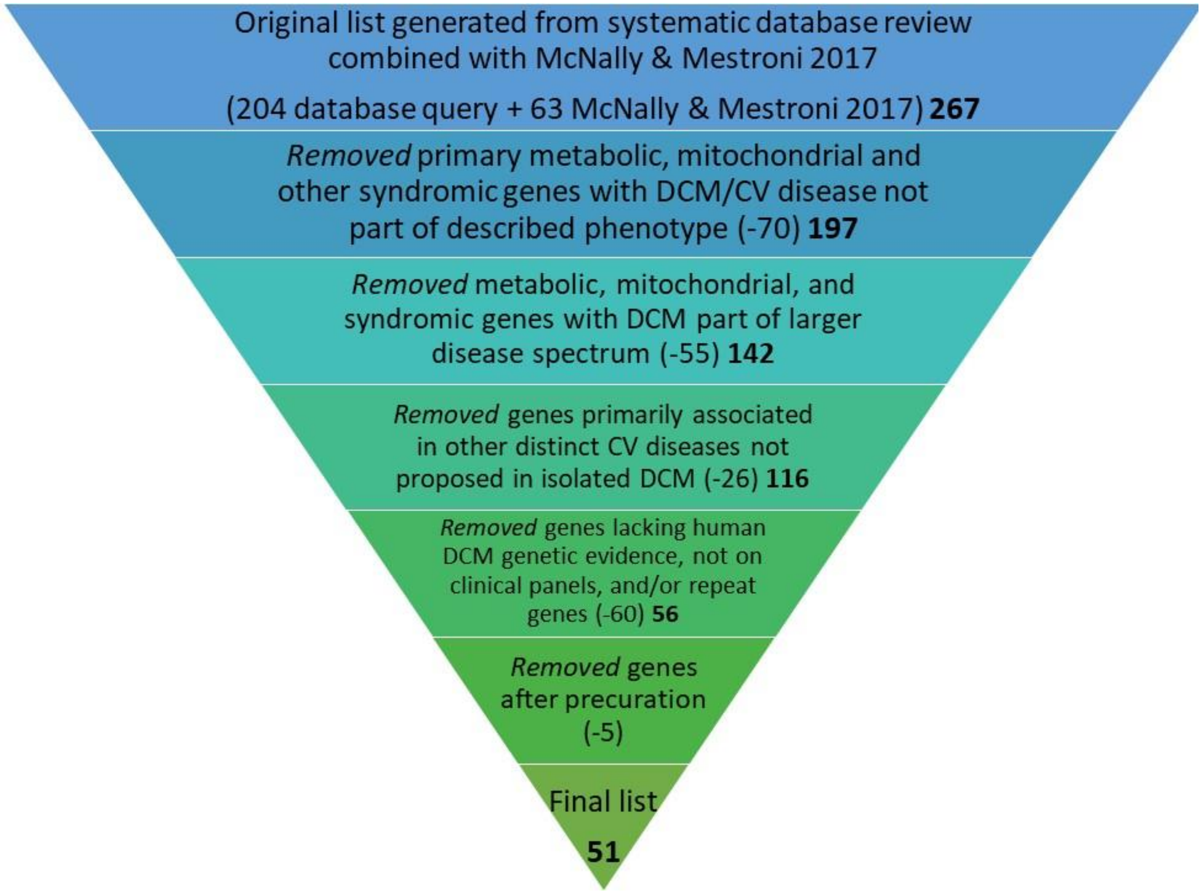


**Figure S1**. **DCM gene list filtering.** An original list of 267 genes, generated by OMIM, Gene, and GenBank database queries combined with a previously published DCM gene list[3], was filtered down to a list of 51 genes indicated in humans with isolated, idiopathic DCM that proceeded through the gene curation process. CV = cardiovascular; DCM = dilated cardiomyopathy.

Supplemental Table 1: Gene List Filtering

| ORIGINAL LIST (OMIM; GENE; GenBank) | McNally 2017 (PMID 28912180) | REMOVED GENES (ALL) | REMOVED- MT/METABOLIC/ SYNDROMIC | REMOVED- NOT IDIOPATHIC DCM (OTHER CM/CV DZ) | REMOVED - NOT IDIOPATHIC DCM (NON-CV DZ/syndrome) | REMOVED - PRECURATION | REMOVED - OTHER | FINAL LIST |
|---|---|---|---|---|---|---|---|---|
| **ABCC9** | **ABCC9** | | | | | | | **ABCC9** |
| ACADVL | | ACADVL | ACADVL | | | | | |
| ACE | | ACE | | | ACE | | | |
| **ACTC1** | **ACTC1** | | | | | | | **ACTC1** |
| **ACTN2** | **ACTN2** | | | | | | | **ACTN2** |
| ADIPOQ | | ADIPOQ | ADIPOQ | | | | | |
| ADORA1 | | ADORA1 | | | ADORA1 | | | |
| ADRA2C | | ADRA2C | | ADRA2C | | | | |
| ADRB1 | | ADRB1 | | ADRB1 | | | | |
| ADRB2 | | ADRB2 | | | ADRB2 | | | |
| | AGL | AGL | AGL | | | | | |
| AGT | | AGT | | | AGT | | | |
| AGTR1 | | AGTR1 | | | AGTR1 | | | |
| ALMS1 | ALMS1 | ALMS1 | ALMS1 | | | | | |
| | ALPK3 | ALPK3 | | | | ALPK3 | | |
| **ANKRD1** | **ANKRD1** | | | | | | | **ANKRD1** |
| ANO5 | | ANO5 | ANO5 | | | | | |
| AR | | AR | | | AR | | | |
| **BAG3** | **BAG3** | | | | | | | **BAG3** |
| BCL2 | | BCL2 | | | | | BCL2 | |
| BMP4 | | BMP4 | | | BMP4 | | | |
| | BRAF | BRAF | BRAF | | | | | |
| BTNL2 | | BTNL2 | | BTLN2 | | | | |
| | CACNA1C | CACNA1C | | CACNA1C | | | | |
| CACNG8 | | CACNG8 | | | | | CACNG8 | |
| | CALR3 | CALR3 | | | | | CALR3 | |
| CASK | | CASK | | | CASK | | | |
| | CASQ2 | CASQ2 | | CASQ2 | | | | |
| CASZ1 | | CASZ1 | | | | | CASZ1 | |
| | CAV3 | CAV3 | | CAV3 | | | | |
| CAVIN4 | (MURC) | CAVIN4/MURC | | | | | REPEAT (MURC) | |
| CD34 | | CD34 | | | | | CD34 | |
| CD40LG | | CD40LG | | | CD40LG | | | |
| CD46 | | CD46 | | | CD46 | | | |
| CDH2 | | CDH2 | CDH2 | | | | | |
| CHGA | | CHGA | | | | | CHGA | |
| | CHRM2 | CHRM2 | | | | | CHRM2 | |
| CHRM3 | | CHRM3 | | | CHRM3 | | | |
| CMD1B | | CMD1B | | | | | CMD1B | |
| CMD1H | | CMD1H | | | | | CMD1H | |
| CMD1K | | CMD1K | | | | | CMD1K | |
| CMD1Q | | CMD1Q | | | | | CMD1Q | |
| | CPT2 | CPT2 | CPT2 | | | | | |
| CRP | | CRP | | | | | CRP | |
| CRYAB | CRYAB | CRYAB | CRYAB | | | | | |
| **CSRP3** | **CSRP3** | | | | | | | **CSRP3** |
| | **CTF1** | | | | | | | **CTF1** |
| CTLA4 | | CTLA4 | | | CTLA4 | | | |
| CTNNA3 | CTNNA3 | CTNNA3 | | CTNNA3 | | | | |
| CTSB | | CTSB | | | CTSB | | | |
| CTSL | | CTSL | | | | | CTSL | |
| CX3CR1 | | CX3CR1 | | CX3CR1 | | | | |
| CXADR | | CXADR | | | CXADR | | | |
| CYP2E1 | | CYP2E1 | | | CYP2E1 | | | |
| DAG1 | | DAG1 | DAG1 | | | | | |
| **DES** | **DES** | | | | | | | **DES** |
| DMD | DMD | DMD | DMD | | | | | |
| DNAJC19 | DNAJC19 | DNAJC19 | DNAJC19 | | | | | |
| | DOLK | DOLK | DOLK | | | | | |
| DSC2 | DSC2 | DSC2 | | | | DSC | | |
| **DSG2** | **DSG2** | | | | | | | **DSG2** |
| | **DSP** | | | | | | | **DSP** |
| | **DTNA** | | | | | | | **DTNA** |
| EDN1 | | EDN1 | | EDN1 | | | | |
| EDNRA | | EDNRA | | | EDNRA | | | |
| EDNRB | | EDNRB | | | EDNRB | | | |
| | EMD | EMD | EMD | | | | | |
| ERBB2 | | ERBB2 | | | ERBB2 | | | |
| ESR1 | | ESR1 | | ESR1 | | | | |
| ESR2 | | ESR2 | | | ESR2 | | | |

| | | | | | | | | |
|---|---|---|---|---|---|---|---|---|
| ESRRB | | ESRRB | | | ESRRB | | | |
| **EYA4** | **EYA4** | | | | | | | **EYA4** |
| FAS | | FAS | | | FAS | | | |
| FBXO32 | | FBXO32 | | | | | FBXO32 | |
| FHOD3 | | FHOD3 | | | | | FHOD3 | |
| | FHL1 | FHL1 | FHL1 | | | | | |
| | FHL2 | FHL2 | | FHL2 | | | | |
| FKRP | FKRP | FKRP | FKRP | | | | | |
| FKTN | FKTN | FKTN | FKTN | | | | | |
| **FLNC** | **FLNC** | | | | | | | **FLNC** |
| FN1 | | FN1 | | | FN1 | | | |
| FOXD4 | | FOXD4 | | | | | FOXD4 | |
| | FRDA/FXN | FRDA/FXN | FRDA/FXN | | | | | |
| | GAA | GAA | GAA | | | | | |
| GATA4 | GATA4 | GATA4 | | | | GATA4 | | |
| GATA6 | GATA6 | GATA6 | | | | GATA6 | | |
| **GATAD1** | **GATAD1** | | | | | | | **GATAD1** |
| GBE1 | | GBE1 | GBE1 | | | | | |
| GDF1 | | GDF1 | | GDF1 | | | | |
| GJA4 | | GJA4 | | | | | GJA4 | |
| | GLA | GLA | GLA | | | | | |
| HAND1 | | HAND1 | | | | | HAND1 | |
| HAVCR1 | | HAVCR1 | | | | | HAVCR1 | |
| | HCN4 | HCN4 | | HCN4 | | | | |
| HFE | HFE | HFE | | HFE | | | | |
| HLA-DQB1 | | HLA-DQB1 | | | HLA-DQB1 | | | |
| HLA-DRB1 | | HLA-DRB1 | | | HLA-DRB1 | | | |
| HLA-DRB4 | | HLA-DRB4 | | | | | HLA-DRB4 | |
| HLA-G | | HLA-G | | | HLA-G | | | |
| | HOPX | HOPX | | | | | HOPX | |
| | HRAS | HRAS | HRAS | | | | | |
| HSPD1 | | HSPD1 | | | HSPD1 | | | |
| IFIH1 | | IFIH1 | | | IFIH1 | | | |
| IFNG | | IFNG | | | IFNG | | | |
| IGF1 | | IGF1 | | | IGF1 | | | |
| IL10 | | IL10 | | | IL10 | | | |
| IL17A | | IL17A | | | | | IL17A | |
| IL17F | | IL17F | | | IL17F | | | |
| IL1B | | IL1B | | | IL1B | | | |
| IL2RA | | IL2RA | | | IL2RA | | | |
| IL6 | | IL6 | | | IL6 | | | |
| | **ILK** | | | | | | | **ILK** |
| ITGA6 | | ITGA6 | | | ITGA6 | | | |
| ITGB1BP2 | | ITGB1BP2 | | | | | ITGB1BP2 | |
| ITGB4 | | ITGB4 | | | ITGB4 | | | |
| ITLN1 | | ITLN1 | | | | | ITLN1 | |
| ITPR2 | | ITPR2 | | | ITPR2 | | | |
| | **JPH2** | | | | | | | **JPH2** |
| | JUP | JUP | | JUP | | | | |
| KCNJ11 | | KCNJ11 | | | KCNJ11 | | | |
| KCNJ12 | | KCNJ12 | | | | | KCNJ12 | |
| KCNJ2 | | KCNJ2 | | KCNJ2 | | | | |
| KCNN3 | | KCNN3 | | | KCNN3 | | | |
| KCNQ1 | KCNQ1 | KCNQ1 | | KCNQ1 | | | | |
| | KRAS | KRAS | KRAS | | | | | |
| LAMA2 | LAMA2 | LAMA2 | LAMA2 | | | | | |
| **LAMA4** | **LAMA4** | | | | | | | **LAMA4** |
| LAMP2 | LAMP2 | LAMP2 | LAMP2 | | | | | |
| LCN2 | | LCN2 | | | | | LCN2 | |
| **LDB3** | **LDB3** | | | | | | | **LDB3** |
| LGALS3 | | LGALS3 | | | | | LGALS3 | |
| **LMNA** | | | | | | | | **LMNA** |
| LRP1 | | LRP1 | | | LRP1 | | | |
| **LRRC10** | **LRRC10** | | | | | | | **LRRC10** |
| LRRC32 | | LRRC32 | | | | | LRRC32 | |
| LTBP4 | | LTBP4 | | | LTBP4 | | | |
| | MAP2K1 | MAP2K1 | MAP2K1 | | | | | |
| | MAP2K2 | MAP2K2 | MAP2K2 | | | | | |
| MAPK14 | | MAPK14 | | | | | MAPK14 | |
| | **MIB1** | | | | | | | **MIB1** |
| MIR199A1 | | MIR199A1 | | | | | MIR199A1 | |
| MIR208A | | MIR208A | | | | | MIR208A | |
| MIR214 | | MIR214 | | | | | MIR214 | |
| MIR451A | | MIR451A | | | | | MIR415A | |
| MLIP | | MLIP | | | | | MLIP | |

| | | | | | | | | |
|---|---|---|---|---|---|---|---|---|
| MMP1 | | MMP1 | | | MMP1 | | | |
| MMP10 | | MMP10 | | | | | MMP10 | |
| MMP14 | | MMP14 | | | MMP14 | | | |
| MMP3 | | MMP3 | | MMP3 | | | | |
| MMP7 | | MMP7 | | | | | MMP7 | |
| MMP9 | | MMP9 | | | MMP9 | | | |
| MT-TI | | MT-TI | MTTI | | | | | |
| | MTND1 | MTND1 | MTND1 | | | | | |
| | MTND5 | MTND5 | MTND5 | | | | | |
| | MTND6 | MTND6 | MTND6 | | | | | |
| | MTTD | MTTD | MTTD | | | | | |
| | MTTG | MTTG | MTTG | | | | | |
| | MTTH | MTTH | MTTH | | | | | |
| | MTTI | MTTI | MTTI | | | | | |
| | MTTK | MTTK | MTTK | | | | | |
| | MTTL1 | MTTL1 | MTTL1 | | | | | |
| | MTTL2 | MTTL2 | MTTL2 | | | | | |
| | MTTM | MTTM | | | MTTM | | | |
| | MTTQ | MTTQ | | | MTTQ | | | |
| | MTTS1 | MTTS1 | | | MTTS1 | | | |
| | MTTS2 | MTTS2 | | | MTTS2 | | | |
| CAVIN4 | MURC | MURC/CAVIN | | | | | MURC/CAVIN | |
| MYBPC1 | | MYBPC1 | | | MYBPC1 | | | |
| **MYBPC3** | **MYBPC3** | | | | | | | **MYBPC3** |
| MYC | | MYC | | | MYC | | | |
| **MYH6** | **MYH6** | | | | | | | **MYH6** |
| **MYH7** | **MYH7** | | | | | | | **MYH7** |
| **MYL2** | **MYL2** | | | | | | | **MYL2** |
| | **MYL3** | | | | | | | **MYL3** |
| | MYLK2 | MYLK2 | | MYLK2 | | | | |
| MYOM1 | MYOM1 | MYOM1 | | MYOM1 | | | | |
| | MYOZ2 | MYOZ2 | | MYOZ2 | | | | |
| **MYPN** | | | | | | | | **MYPN** |
| NAMPT | | NAMPT | | | | | NAMPT | |
| NDUFV1 | | NDUFV1 | | | NDUFV1 | | | |
| **NEBL** | **NEBL** | | | | | | | **NEBL** |
| **NEXN** | **NEXN** | | | | | | | **NEXN** |
| NFKB1 | NFKB1 | NFKB1 | | | NFKB1 | | | |
| | **NKX2-5** | | | | | | | **NKX2-5** |
| NLRP3 | | NLRP3 | | | NLRP3 | | | |
| NOS3 | | NOS3 | | | NOS3 | | | |
| **NPPA** | **NPPA** | | | | | | | **NPPA** |
| NPPB | | NPPB | | | | | NPPB | |
| NPPC | | NPPC | | | NPPC | | | |
| NPR2 | | NPR2 | | | NPR2 | | | |
| NR3C2 | | NR3C2 | | | NR3C2 | | | |
| | NRAS | NRAS | NRAS | | | | | |
| **OBSCN** | | | | | | | | OBSCN |
| OSM | | OSM | | | | | OSM | |
| PDCD1 | | PDCD1 | | | PDCD1 | | | |
| PDE2A | | PDE2A | | | | | PDE2A | |
| PDE3A | | PDE3A | | | PDE3A | | | |
| | **PDLIM3** | | | | | | | **PDLIM3** |
| PGM1 | | PGM1 | PGM1 | | | | | |
| | **PKP2** | | | | | | | **PKP2** |
| PLEC1 | PLEC1 | PLEC1 | PLEC | | | | | |
| **PLEKHM2** | **PLEKHM2** | | | | | | | **PLEKHM2** |
| **PLN** | **PLN** | | | | | | | **PLN** |
| PNPLA2 | | PNPLA2 | PNPLA2 | | | | | |
| POLG | | POLG | POLG | | | | | |
| PPP1R13L | | PPP1R13L | | | | | PPP1R13L | |
| **PRDM16** | **PRDM16** | | | | | | | **PRDM16** |
| | PRKAG2 | PRKAG2 | | PRKAG2 | | | | |
| **PSEN1** | | | | | | | | **PSEN1** |
| **PSEN2** | | | | | | | | **PSEN2** |
| | PTPN11 | PTPN11 | PTPN11 | | | | | |
| RAF1 | RAF1 | RAF1 | RAF1 (Noonan/Rasopathy) | | | | | |
| RARRES2 | | RARRES2 | | | | | RARRES2 | |
| **RBM20** | **RBM20** | | | | | | | **RBM20** |
| | RIT1 | RIT1 | RIT1 (noonan/rasopathy) | | | | | |
| RYR2 | RYR2 | RYR2 | | RYR2 | | | | |

| | | | | | | | | |
|---|---|---|---|---|---|---|---|---|
| **SCN5A** | **SCN5A** | | | | | | | **SCN5A** |
| SDHA | SDHA | | SDHA | | | SDHA | | |
| SERPINE1 | | SERPINE1 | | | SERPINE1 | | | |
| | SGCA | SGCA | SGCA | | | | | |
| | SGCB | SGCB | SGCB | | | | | |
| **SGCD** | **SGCD** | | | | | | | **SGCD** |
| | SGCG | SGCG | SGCG | | | | | |
| SGK1 | | SGK1 | | | | | SGK1 | |
| | SLC22A5 | SLC22A5 | SLC22A5 | | | | | |
| SLC25A5P8 | | SLC25A5P8 | | | | | SLC25A5P8 | |
| SOD2 | SOD2 | SOD2 | | | | | SOD2 | |
| | SOS1 | SOS1 | SOS1 | | | | | |
| SPEG | | SPEG | SPEG | | | | | |
| SRF | | SRF | | | | | SRF | |
| STAT3 | | STAT3 | | | STAT3 | | | |
| SYNE1 | | SYNE1 | SYNE3 | | | | | |
| | SYNM | SYNM | | | | | SYNM | |
| TAX1BP3 | | TAX1BP3 | | | | | TAX1BP3 | |
| TAZ | TAZ | TAZ | TAZ | | | | | |
| **TBX20** | **TBX20** | | | | | | | **TBX20** |
| TBX5 | | TBX5 | | | TBX5 | | | |
| **TCAP** | **TCAP** | | | | | | | **TCAP** |
| TFAP2A | | TFAP2A | | | TFAP2A | | | |
| TGFB1 | | TGFB1 | | | TGFB1 | | | |
| | TGFB3 | TGFB3 | | TGFB3 | | | | |
| TIMP1 | | TIMP1 | | | | | TIMP1 | |
| TIMP4 | | TIMP4 | | | | | TIMP4 | |
| | TMEM43 | TMEM43 | TMEM43 | | | | | |
| TMPO | | TMPO | | | | | TMPO | |
| TNC | | TNC | | | TNC | | | |
| TNF | | TNF | | | TNF | | | |
| TNFRSF11B | | TNFRSF11B | | | TNFRSF11B | | | |
| TNFRSF12A | | TNFRSF12A | | | | | TNFRSF12A | |
| TNFSF10 | | TNFSF10 | | | | | TITNFSF10 | |
| TNFSF12 | | TNFSF12 | | | | | TNFSF1 | |
| **TNNC1** | **TNNC1** | | | | | | | **TNNC1** |
| **TNNI3** | **TNNI3** | | | | | | | **TNNI3** |
| **TNNI3K** | **TNNI3K** | | | | | | | **TNNI3K** |
| **TNNT2** | **TNNT2** | | | | | | | **TNNT2** |
| TP53 | | TP53 | | | TP53 | | | |
| **TPM1** | **TPM1** | | | | | | | **TPM1** |
| | TRDN | TRDN | | TRDN | | | | |
| TRPV2 | | TRPV2 | | | | | TRPV2 | |
| **TTN** | TTN | | | | | | | **TTN** |
| | TTR | TTR | | TTR | | | | |
| TWIST1 | | TWIST1 | | | TWIST | | | |
| TXNRD2 | TXNRD2 | TXNRD2 | TXNRD2 | | | | | |
| **VCL** | **VCL** | | | | | | | **VCL** |
| VEGFA | | VEGFA | | | VEGFA | | | |
| | | | | | | | REPEAT - SEE | |
| ZASP | | ZASP | | | | | LDB3 | |
| ZBTB17 | ZBTB17 | ZBTB17 | | | | | ZBTB17 | |
| **TOTAL: 204** | **TOTAL: 63** (125-62) | **TOTAL: 216** | **TOTAL: -55** | **TOTAL: -26** | **TOTAL: -70** | **TOTAL: -5** | **TOTAL: -60** | **TOTAL: 51** |
| **TOTAL BEFORE REMOVED: 267** | | **TOTAL AFTER REMOVED: 51** | **TOTAL OF ALL REMOVED CATEGORIES: 216** | | | | | **TOTAL FINAL LIST: 51** |

Supplemental Table 2: DCM Panel Composition

| DCM Classification | Curated Genes | USA: Invitae | USA: EGL Genetic Diagnostics | USA: Prevention Genetics | USA: Cincinnati's Childrens Hospital | USA: Ambry Genetics | Finland: Blueprint Genetics | USA: Fulgent Genetics | USA: Washington University | Spain: Health in Code | USA: Knight Diagnostic Laboratories | Spain: CGC Genetics | USA: Gene Dx | Germany: CeGaT GmbH | Germany: MGZ - Medical Genetics Center | Canada: LifeLabs Genetics | USA: Sema4 | Panel Totals | % Panels Present |
|---|---|---|---|---|---|---|---|---|---|---|---|---|---|---|---|---|---|---|---|
| Definitive | BAG3 | 1 | 1 | 1 | 1 | 1 | 1 | 1 | 1 | 1 | 1 | 1 | 1 | 1 | 1 | 1 | 1 | 16 | 100.0% |
| | DES | 1 | 1 | 1 | 1 | 1 | 1 | 1 | 1 | 1 | 1 | 1 | 1 | 1 | 1 | 1 | 1 | 16 | 100.0% |
| | LMNA | 1 | 1 | 1 | 1 | 1 | 1 | 1 | 1 | 1 | 1 | 1 | 1 | 1 | 1 | 1 | 1 | 16 | 100.0% |
| | MYH7 | 1 | 1 | 1 | 1 | 1 | 1 | 1 | 1 | 1 | 1 | 1 | 1 | 1 | 1 | 1 | 1 | 16 | 100.0% |
| | PLN | 1 | 1 | 1 | 1 | 1 | 1 | 1 | 1 | 1 | 1 | 1 | 1 | 1 | 1 | 1 | 1 | 16 | 100.0% |
| | RBM20 | 1 | 1 | 1 | 1 | 1 | 1 | 1 | 1 | 1 | 1 | 1 | 1 | 1 | 1 | 1 | 1 | 16 | 100.0% |
| | SCN5A | 1 | 1 | 1 | 1 | 1 | 1 | 1 | 1 | 1 | 1 | 1 | 1 | 1 | 1 | 1 | 1 | 16 | 100.0% |
| | TTN | 1 | 1 | 1 | 1 | 1 | 1 | 1 | 1 | 1 | 1 | 1 | 1 | 1 | 1 | 1 | 1 | 16 | 100.0% |
| | TNNC1 | 1 | 1 | 1 | 1 | 1 | 1 | 1 | 1 | 1 | 0 | 1 | 1 | 1 | 1 | 1 | 1 | 15 | 93.8% |
| | TNNT2 | 1 | 1 | 1 | 1 | 1 | 1 | 1 | 1 | 1 | 0 | 1 | 1 | 1 | 1 | 1 | 1 | 15 | 93.8% |
| | FLNC | 1 | 0 | 1 | 1 | 1 | 1 | 1 | 1 | 1 | 0 | 1 | 1 | 1 | 0 | 0 | 1 | 12 | 75.0% |
| | | | | | | | | | | | | | | | | | | 0 | 0.0% |
| Strong | DSP | 1 | 1 | 1 | 1 | 0 | 1 | 1 | 0 | 1 | 1 | 1 | 1 | 1 | 1 | 1 | 1 | 14 | 87.5% |
| | | | | | | | | | | | | | | | | | | 0 | 0.0% |
| Moderate | ACTC1 | 1 | 1 | 1 | 1 | 1 | 1 | 1 | 1 | 1 | 1 | 1 | 1 | 1 | 1 | 1 | 1 | 16 | 100.0% |
| | TNNI3 | 1 | 1 | 1 | 1 | 1 | 1 | 1 | 1 | 1 | 1 | 1 | 1 | 1 | 1 | 1 | 1 | 16 | 100.0% |
| | TPM1 | 1 | 1 | 1 | 1 | 1 | 1 | 1 | 1 | 1 | 1 | 1 | 1 | 1 | 1 | 1 | 1 | 16 | 100.0% |
| | VCL | 1 | 1 | 1 | 1 | 1 | 1 | 1 | 1 | 1 | 1 | 1 | 1 | 1 | 1 | 1 | 1 | 16 | 100.0% |
| | ACTN2 | 0 | 1 | 1 | 1 | 1 | 1 | 1 | 1 | 1 | 1 | 1 | 1 | 1 | 1 | 1 | 1 | 15 | 93.8% |
| | NEXN | 1 | 1 | 1 | 1 | 1 | 0 | 1 | 1 | 1 | 1 | 1 | 1 | 1 | 1 | 1 | 1 | 15 | 93.8% |
| | JPH2 | 0 | 0 | 0 | 1 | 0 | 1 | 1 | 0 | 0 | 0 | 0 | 0 | 1 | 0 | 0 | 0 | 4 | 25.0% |
| | | | | | | | | | | | | | | | | | | 0 | 0.0% |
| Limited | ABCC9 | 1 | 1 | 1 | 1 | 1 | 1 | 1 | 1 | 1 | 1 | 1 | 1 | 1 | 1 | 1 | 1 | 16 | 100.0% |
| | LDB3 | 1 | 1 | 1 | 1 | 1 | 1 | 1 | 1 | 1 | 1 | 1 | 1 | 1 | 1 | 1 | 1 | 16 | 100.0% |
| | MYBPC3 | 1 | 1 | 1 | 1 | 1 | 1 | 1 | 1 | 1 | 1 | 1 | 1 | 1 | 1 | 1 | 1 | 16 | 100.0% |
| | MYH6 | 1 | 1 | 1 | 1 | 1 | 1 | 1 | 1 | 1 | 1 | 1 | 1 | 1 | 1 | 1 | 1 | 16 | 100.0% |
| | TCAP | 1 | 1 | 1 | 1 | 1 | 1 | 1 | 1 | 1 | 1 | 1 | 1 | 1 | 1 | 1 | 1 | 16 | 100.0% |
| | ANKRD1 | 1 | 1 | 1 | 1 | 1 | 0 | 1 | 1 | 1 | 1 | 1 | 1 | 1 | 1 | 1 | 1 | 15 | 93.8% |
| | CSRP3 | 1 | 1 | 1 | 1 | 1 | 0 | 1 | 1 | 1 | 1 | 1 | 1 | 1 | 1 | 1 | 1 | 15 | 93.8% |
| | LAMA4 | 1 | 1 | 1 | 1 | 1 | 1 | 1 | 0 | 1 | 1 | 1 | 1 | 1 | 1 | 1 | 1 | 15 | 93.8% |
| | MYPN | 1 | 1 | 1 | 1 | 1 | 0 | 1 | 1 | 1 | 1 | 1 | 1 | 1 | 1 | 1 | 1 | 15 | 93.8% |
| | DSG2 | 1 | 1 | 1 | 1 | 0 | 1 | 1 | 0 | 1 | 1 | 1 | 1 | 1 | 1 | 1 | 1 | 14 | 87.5% |
| | GATAD1 | 1 | 1 | 1 | 1 | 0 | 0 | 1 | 1 | 1 | 1 | 1 | 1 | 1 | 1 | 1 | 1 | 14 | 87.5% |
| | ILK | 1 | 0 | 1 | 1 | 0 | 1 | 1 | 1 | 1 | 1 | 1 | 1 | 1 | 1 | 1 | 1 | 14 | 87.5% |
| | NEBL | 1 | 1 | 1 | 1 | 0 | 0 | 1 | 1 | 1 | 1 | 1 | 1 | 1 | 0 | 1 | 1 | 13 | 81.3% |
| | PRDM16 | 1 | 1 | 1 | 1 | 0 | 1 | 1 | 0 | 1 | 0 | 1 | 1 | 1 | 1 | 1 | 1 | 13 | 81.3% |
| | SGCD | 0 | 1 | 1 | 1 | 0 | 0 | 1 | 1 | 1 | 1 | 1 | 1 | 1 | 1 | 1 | 1 | 13 | 81.3% |
| | EYA4 | 1 | 0 | 1 | 1 | 0 | 0 | 1 | 1 | 1 | 1 | 1 | 0 | 0 | 1 | 1 | 1 | 11 | 68.8% |
| | NKX2-5 | 1 | 0 | 1 | 1 | 1 | 1 | 1 | 1 | 1 | 0 | 1 | 1 | 0 | 0 | 0 | 0 | 10 | 62.5% |
| | TBX20 | 0 | 0 | 0 | 1 | 1 | 1 | 1 | 1 | 1 | 0 | 0 | 1 | 0 | 1 | 1 | 0 | 9 | 56.3% |
| | DTNA | 0 | 0 | 0 | 1 | 0 | 0 | 1 | 0 | 1 | 0 | 0 | 1 | 1 | 0 | 0 | 0 | 5 | 31.3% |
| | MYL2 | 0 | 0 | 0 | 1 | 0 | 0 | 1 | 0 | 1 | 0 | 1 | 0 | 1 | 0 | 0 | 0 | 5 | 31.3% |
| | CTF1 | 1 | 0 | 0 | 0 | 0 | 0 | 1 | 1 | 0 | 0 | 1 | 0 | 0 | 0 | 0 | 0 | 4 | 25.0% |
| | PLEKHM2 | 1 | 0 | 0 | 0 | 0 | 1 | 0 | 1 | 0 | 0 | 0 | 0 | 0 | 0 | 0 | 0 | 3 | 18.8% |
| | PSEN2 | 0 | 0 | 0 | 0 | 0 | 0 | 0 | 0 | 1 | 0 | 1 | 0 | 0 | 1 | 0 | 0 | 3 | 18.8% |
| | TNNI3K | 0 | 0 | 0 | 0 | 1 | 1 | 0 | 0 | 1 | 0 | 0 | 0 | 0 | 0 | 0 | 0 | 3 | 18.8% |
| | OBSCN | 0 | 0 | 0 | 0 | 0 | 0 | 0 | 0 | 1 | 0 | 0 | 0 | 0 | 0 | 0 | 1 | 2 | 12.5% |
| | | | | | | | | | | | | | | | | | | 0 | 0.0% |
| Disputed | PDLIM3 | 1 | 1 | 1 | 1 | 0 | 0 | 1 | 1 | 1 | 1 | 1 | 0 | 1 | 1 | 1 | 0 | 12 | 75.0% |

| | | | | | | | | | | | | | | | | | | | |
|---|---|---|---|---|---|---|---|---|---|---|---|---|---|---|---|---|---|---|---|
| | PKP2 | 1 | 0 | 1 | 1 | 0 | 1 | 1 | 0 | 1 | 1 | 1 | 0 | 1 | 1 | 1 | 1 | 12 | 75.0% |
| | MYL3 | 0 | 0 | 0 | 1 | 0 | 0 | 1 | 0 | 1 | 0 | 1 | 0 | 1 | 0 | 0 | 0 | 5 | 31.3% |
| | PSEN1 | 0 | 0 | 0 | 0 | 0 | 0 | 0 | 0 | 1 | 0 | 1 | 0 | 0 | 1 | 0 | 0 | 3 | 18.8% |
| | | | | | | | | | | | | | | | | | | 0 | 0.0% |
| No Known Dise | MIB1 | 0 | 0 | 1 | 1 | 0 | 0 | 1 | 0 | 1 | 0 | 0 | 1 | 0 | 0 | 0 | 0 | 5 | 31.3% |
| | LRRC10 | 1 | 0 | 0 | 1 | 0 | 1 | 0 | 1 | 0 | 0 | 0 | 1 | 0 | 0 | 0 | 0 | 5 | 31.3% |
| | NPPA | 1 | 0 | 0 | 0 | 0 | 0 | 1 | 1 | 0 | 0 | 1 | 0 | 0 | 0 | 0 | 0 | 4 | 25.0% |
